# Enough evidence and other endings: a descriptive study of stable Cochrane systematic reviews in 2019

**DOI:** 10.1101/19013912

**Authors:** Hilda Bastian, Lars G. Hemkens

**Affiliations:** doctoral candidate, Bond University, Australia; Basel Institute for Clinical Epidemiology and Biostatistics, University Hospital Basel, Switzerland

## Abstract

**Background:** From 2006 to 2019, Cochrane reviews could be designated “stable” if they were not being updated but highly likely to be current. This provides an opportunity to observe practice in ending systematic reviewing and what is regarded as enough evidence.

**Methods:** We identified Cochrane reviews designated stable in 2013 and 2019 and reasons for this designation. For those with conclusions stated to be so firm that new evidence is unlikely to change them, we assessed conclusions, strength of evidence ratings, and recommendations for further research. We assessed the fate of the 2013 stable reviews. We also estimated usage of formal analytic methods to determine when there is enough evidence in protocols for Cochrane reviews.

**Results:** Cochrane reviews were rarely designated stable. In 2019, there were 507 stable Cochrane reviews (6.6% of 7,645 non-withdrawn reviews). The most common reasons related to no, little, or infrequent research activity expected (331 of 505; 65.5%). Only 39 reviews were stable because of firm conclusions unlikely to be changed by new evidence (7.7%), but that declaration was mostly not supported by judgments made in the review about strength of evidence and implications for research. Among the 180 reviews stable in 2013, 16 reverted to normal status (8.9%), with 2 of those changing conclusions because of new studies. Few Cochrane protocols specified an analytic method for determining when there was enough evidence to stop updating the review (116 of 2,415; 4.8%).

**Conclusion:** Cochrane reviews were more likely to end because important future primary research activity was believed to be unlikely, than because there was enough evidence. Judgments about the strength of evidence and need for research were often inconsistent with the declaration that conclusions were unlikely to change. The inconsistencies underscore the need for reliable analytic methods to support decision-making about the conclusiveness of evidence.

## Background

The question of when there is enough evidence to be certain about effects in health is a critical one: both premature and overdue certainty can do damage. In 1992, Antman and colleagues demonstrated that harmful clinical advice and initiation of redundant clinical trials could continue long past the time a question has been definitively answered. (1) They argued that keeping on top of trial results with systematic reviews could help reduce this problem, a point of view reiterated by Chalmers and Glasziou in a 2009 paper on avoidable research waste. (2)

Systematic reviews involve searching for studies on a question and synthesising the findings, using explicit formal methods. (3) They can be outdated by subsequent studies, sometimes quite quickly. (4) Systematic reviews of clinical trials therefore need to be monitored and updated as long as a critical question about effectiveness or safety remains. This was made feasible on a large scale by developments in information technology and publishing, and enabled the establishment of the Cochrane Collaboration in 1993. (5) The Cochrane Collaboration developed a reviewer and methodologist community around the production and updating of systematic reviews, underpinned by a clinical trials register and dissemination through its own journal, the *Cochrane Database of Systematic Reviews* (CDSR). (6,7) The Cochrane community also developed a range of technical and social mechanisms, and played an often-pioneering role in establishing and evaluating scientific methodology. (8)

The Collaboration had an initial goal of continual updating and at least one update a year, moving to a default update interval of two years in 2000, (9) and dropping the routine updating expectation by 2019. (10) Attempting to keep up with evidence in health like this raises a variety of complex issues. (11) Methods for updating systematic reviews became a widespread concern among systematic reviewers and users of systematic reviews, as well as an area of study. (3,12–14) At some point, updating a systematic review can become redundant, but there is a risk in misjudging this.

In the wake of a mega-trial that overturned the results of a meta-analysis in 1995, Egger and Smith argued “several medium sized trials of high quality seem necessary to render results trustworthy”. (15) Pogue and Yusuf developed a method using optimal information size and cumulative meta-analysis, advocating for its use for prospective determination and monitoring of what would be enough evidence in systematic review protocols, just as for clinical trials. (16,17) Egger et al identified limitations in this method, and it never crossed over into practice. (18,19)

Wetterslev et al built on the Pogue/Yusuf method in 2008, incorporating consideration of heterogeneity, with a method they called trial sequential analysis with cumulative meta-analysis. (20–22) Cochrane’s 2019 guidance to reviewers discourages the use of these methods for updating reviews, unless it is prospectively included in the protocol and used as a secondary analysis only, or in a prospective meta-analysis of a defined group of trials. (10) A 2016 consensus report shows that there is still no well-validated methodology in routine use in systematic reviewing, however, that can reliably show when we have reached, or passed, the point of “enough evidence”. (14)

Up until 2006, there were two status options for Cochrane reviews: “normal” (active reviews, ordinarily with an interval of two years until update was due), and withdrawn (the review is retracted). In 2006, the Collaboration’s governing body decided to add a third option, designating a review as “stable”, in the following software release for Cochrane reviews. (23) This status was codified in the 2008 version of the organisation’s handbook for systematic reviewers, (24) defining a stable review as one that is no longer updated but “highly likely to maintain its current relevance for the foreseeable future (measured in years rather than months)”. This status was to be reviewed periodically, and two uses were specified:

- “The intervention is superseded (bearing in mind that Cochrane reviews should be internationally relevant);
- The conclusion is so certain that the addition of new information will not change it, and there are no foreseeable adverse effects of the intervention”.

In 2008, the Grading of Recommendations, Assessment, Development and Evaluation (GRADE) Group introduced a categorisation of “high quality” in assessing the strength of evidence, defined as “Further research is very unlikely to change our confidence in the estimate of effect”, and this was soon incorporated into Cochrane review summary of findings tables. (24,25) However, the GRADE Group moved away from this conceptualisation by 2017, in favour of “high certainty of evidence”. (26) The GRADE Handbook also recommends using an optimal or required information size calculation to assess the adequacy of precision of an estimate of effect, and that was incorporated in the 2019 edition of the Cochrane Handbook. (10,27)

Cochrane retired the status “stable” in 2019, and a new updating status of “no longer being updated” adopted, with different criteria for use. (10,28) The pool of systematic reviews that have carried the designation “stable” provides an opportunity to study a critical stage in the life cycle of systematic reviews, and when some systematic reviewers believe there is enough evidence. The aims of this study were to assess the extent of usage of stable status, review the fate of reviews designated stable in 2013, and categorise reported reasons for cessation of systematic review updating. We also aimed to describe the reviews with conclusions stated to be unlikely to change with results of new studies, and estimate the extent to which Cochrane reviews were using analytic methods over and above meta-analysis to determine when there was enough evidence.

## Methods

### Study aim 1: assessing the extent of usage of stable status

The identification numbers of all Cochrane systematic reviews designated as stable in the March 2012 and February 2013 issues of the *CDSR* had been identified from the journal’s encoded version (XML markup language), as well as the number of non-withdrawn published Cochrane reviews. Each review’s identification number was recorded, as well as the year it was designated stable according to the published “What’s New” section of the review.

In 2019, we identified the *CDSR* reviews designated stable up to 20 August via the advanced search option in Archie (the internal Cochrane contributors’ database on Cochrane reviews and other documents). (29,30) Identification number, title, and status, were collected, as well as the Cochrane Review Group (CRG) responsible for the review. CRGs are the editorial groups responsible for reviews in specific topic areas. We also collected two fields related to the review’s updating status (“rationale” and “explanation”), which had been introduced in a new Updating Classification System in 2016. (28) Cases where a review had both stable and retracted (“withdrawn”) status were excluded. Reviews that were stable in 2013 and still described as stable in the *CDSR* were included in the study, even if they were not among the reviews declared stable downloaded from Archie.

The total number of Cochrane reviews on 20 August, excluding those that were withdrawn, was obtained from the Cochrane Editorial Unit. A full listing of the CRGs in 2019 was compiled from The Cochrane Library website in March 2019. (31) The reviews of a CRG that no longer exists (HIV/AIDS) were merged with those of the CRG now responsible for that subject area (Infectious Diseases).

### Study aim 2: reviewing the fate of reviews designated stable in 2013

We compared the list of stable reviews from 2013 with those in 2019, identifying those that were no longer designated stable. Data on the current status of those that were no longer designated stable were collected from the *CDSR*, and events reported in the “What’s New” table since 2013 were summarized by one author (HB). Each review’s status was categorised as normal, stable, or withdrawn. These cases were evaluated by both authors.

### Study aim 3: categorising reported reasons for cessation of systematic reviewing

In 2012 and 2013, both authors had reviewed the reasons for the designation given in the “What’s New” section, assigning categories which were developed and agreed on iteratively. Where reasons were not given in the “What’s New” section, the abstract, discussion, and conclusion sections were reviewed. Differences in category assignment were resolved by discussion.

We modified and added to our categories from 2013 when reviews did not fit an existing category. We also used the information in the unpublished fields for update classification in Archie to supplement the information published in the reviews for our categorisation.

We assessed the currency of our 2013 categorisations by checking them against the unpublished “rationale”. When these matched, we retained our original category without further review.

Where the internal “rationale” and “explanation” for a review were unambiguously consistent, and unambiguously matched one of our categories, we assigned that category and undertook no further analysis, unless it was a review we had categorised as having a firm conclusion in 2013. All other cases were initially reviewed by one author (HB), who extracted data on reasons for the designation from the “What’s New” section of that review on the *CDSR* and assigned a category to the review. A random sample of 70 of these were independently assigned a category by the second author (LGH), and differences were resolved by discussion. The second author also reviewed all cases assigned as reaching firm conclusions, and differences were resolved by discussion. Of the final sample of included stable reviews, 42% were assessed by both authors, 27% by one author (HB), and 31% were based on Cochrane classifications alone (S5 File).

### Study aim 4: describing reviews with firm conclusions unlikely to change with new studies

To explore this category of reviews, we categorised the main conclusions of these reviews, resolving differences by discussion. Authors’ judgments in two other parts of Cochrane reviews are directly relevant to aspects of a firm conclusion, and could be expected to support the review’s conclusiveness. The first is the authors’ conclusion on the implications of their findings for research, and the other is the judgment on strength of evidence. We categorised the reviews’ section on implications for future research, resolving differences by discussion.

In addition, one author (HB) collected the highest GRADE rating for certainty of evidence in the summary of findings (SoF) table. Where there was no summary of findings table, the description of evidence quality or certainty in the abstract, results, or discussion sections of the review was collected. When the only evidence rating was at the individual study level, the rating for the best-rated study was collected. To determine whether these reviews reported used specific formal analytic method in addition to meta-analysis to reach their determination of enough evidence, the methods, results, and discussion sections were also reviewed by one author (HB).

### Study aim 5: estimating the extent of usage of formal analytic methods to determine

#### when there is enough evidence in protocols

To estimate the potential usage of analytic methods for analysing whether there is enough evidence above and beyond meta-analysis, a full text search of all protocols in *CDSR* was done for the phrases “trial sequential analysis”, “value of information”, “optimal information size”, or “required information size” on 29 September 2019. Protocols were screened by one author (HB) to identify those that included an analytic method for determining when there would be enough evidence. The number of protocols in the *CDSR* was recorded.

#### Data management and analysis

Data were collected in Excel and analysed using RStudio 1.1463 running R 3.5.2, (32,33) using tidyverse and reshape2 packages. (34,35) Summary statistics were used to describe the cohort. Data for this project, including analytic code, are also deposited at GitHub. (36)

## Results

### Usage of stable status

We identified 507 reviews classified stable among 7,645 non-withdrawn reviews (6.6%) in August 2017 (Figure 1, S3 File, S4 File). In February 2013 there had been 180 Cochrane reviews classified stable, which were 3.5% of all 5,137 non-withdrawn Cochrane reviews.

**Figure 1.**
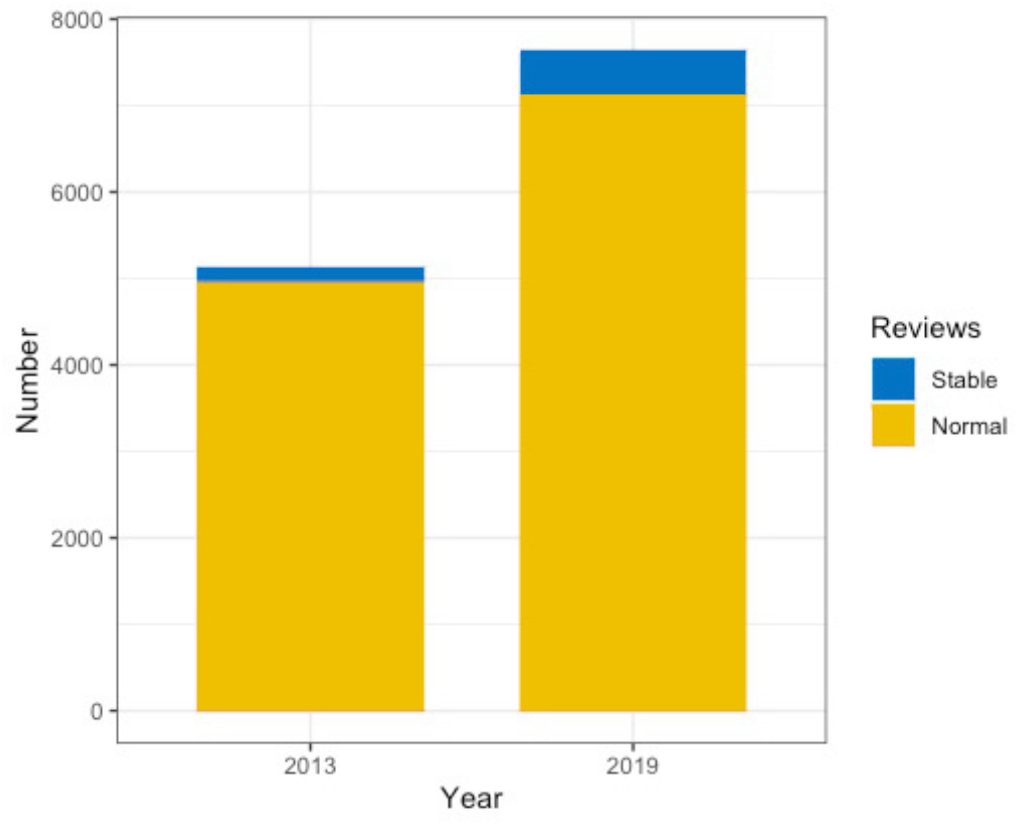
The number of stable reviews among non-withdrawn Cochrane reviews, 2013 and 2019.

There was an increase in the proportion of reviews designated stable between 2013 and 2019. This was in large part due to a single Cochrane Review Group (CRG). Table 1 shows the breakdown of stable reviews across CRGs in 2019, showing that they are not distributed evenly across the 53 groups and most are designated stable by a few groups.

**Table 1.** Stable reviews across Cochrane Review Groups (CRGs).

|  | 0 stable reviews |  | 1-10 stable reviews |  | 11-20 stable reviews |  | >20 stable reviews* |  |
| --- | --- | --- | --- | --- | --- | --- | --- | --- |
|  | No. | % | No. | % | No. | % | No. | % |
| Number and proportion of CRGs by level of volume of stable reviews (n = 53) | 20 | 37.7 | 23 | 43.4 | 4 | 7.5 | 6 | 11.3 |
| Number and proportion of stable reviews in CRGs by level of volume of stable reviews (n = 507) | - | - | 79 | 15.6 | 52 | 10.3 | 376 | 74.2 |

\* Range of 24 to 219

The number of stable reviews per CRG ranged from 0 to 219, with a median of 1 (IQR 7). Of the six CRGs responsible for over 20 stable reviews, three CRGs were responsible for 59.2% of all stable reviews, with a single CRG designating 219 reviews stable (43.2% of all stable reviews).

### Fate of reviews designated stable in 2013

Most of the 180 reviews with stable status in 2013 were still designated stable in 2019 (n = 159, 88.3%), but 16 reverted to normal status (8.9%) and five were withdrawn (2.8%) (Figure 2). One was withdrawn because it was no longer a priority for the editorial group, and the others were being replaced by one or more new reviews or protocols.

**Figure 2.**
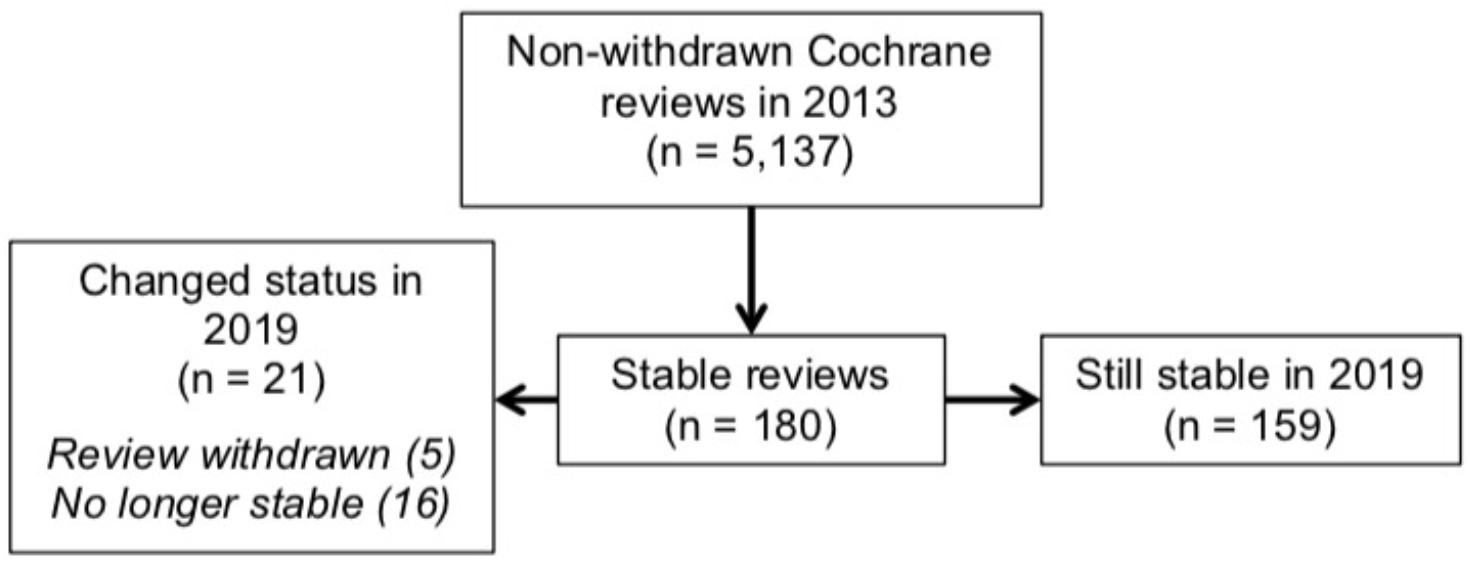
Status in 2019 of stable reviews from February 2013.

Most stable reviews that reverted to normal status did not have new included studies (11 of 16; 68.8%) (Table 2). Two of the reviews had been declared stable in 2013 because of firm conclusions judged unlikely to be changed by new evidence. In one case, however, it was because new trials had been found and an update of the review was in progress. In the other, it was because of a reader’s criticism that the firm conclusion was unjustified.

**Table 2.**
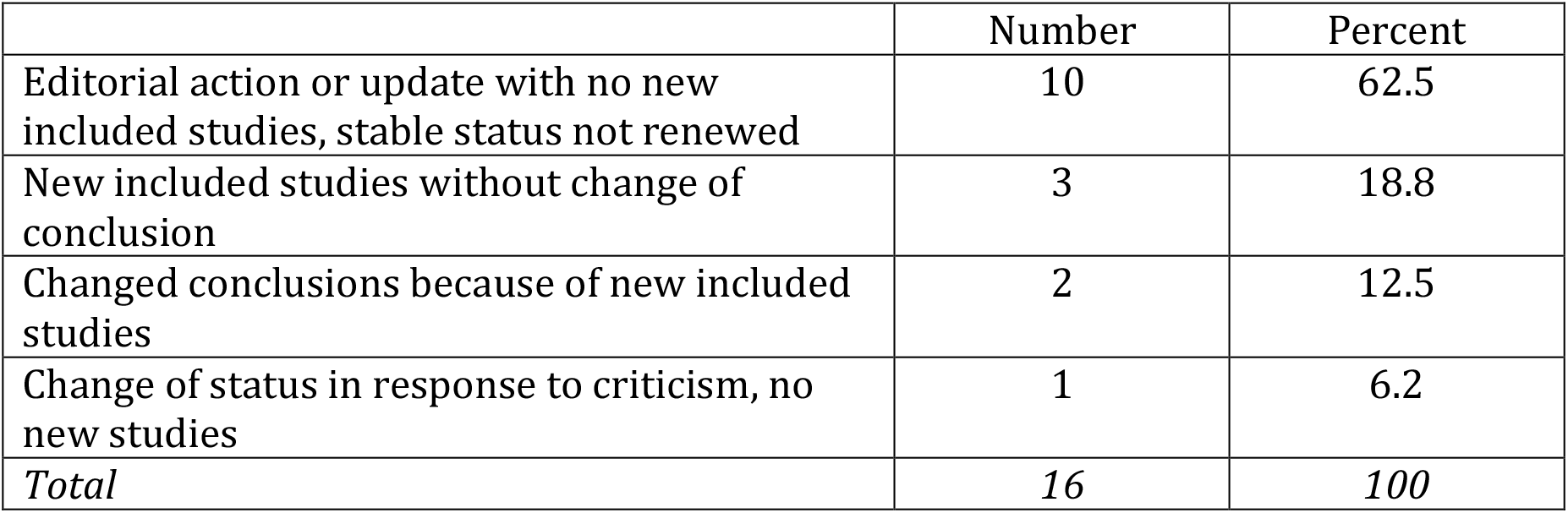
Formerly stable reviews reverted to normal status (n=16).

|  | Number | Percent |
| --- | --- | --- |
| Editorial action or update with no new included studies, stable status not renewed | 10 | 62.5 |
| New included studies without change of conclusion | 3 | 18.8 |
| Changed conclusions because of new included studies | 2 | 12.5 |
| Change of status in response to criticism, no new studies | 1 | 6.2 |
| *Total* | *16* | *100* |

### Reasons for reviews being designated stable

We agreed on eight categories for the reasons reported for declaring reviews stable. In Figure 3, we summarise these reasons and group them into those with an alternative schedule for updating, those where updates have ceased, and those where it appears that updating was never intended.

**Figure 3.**
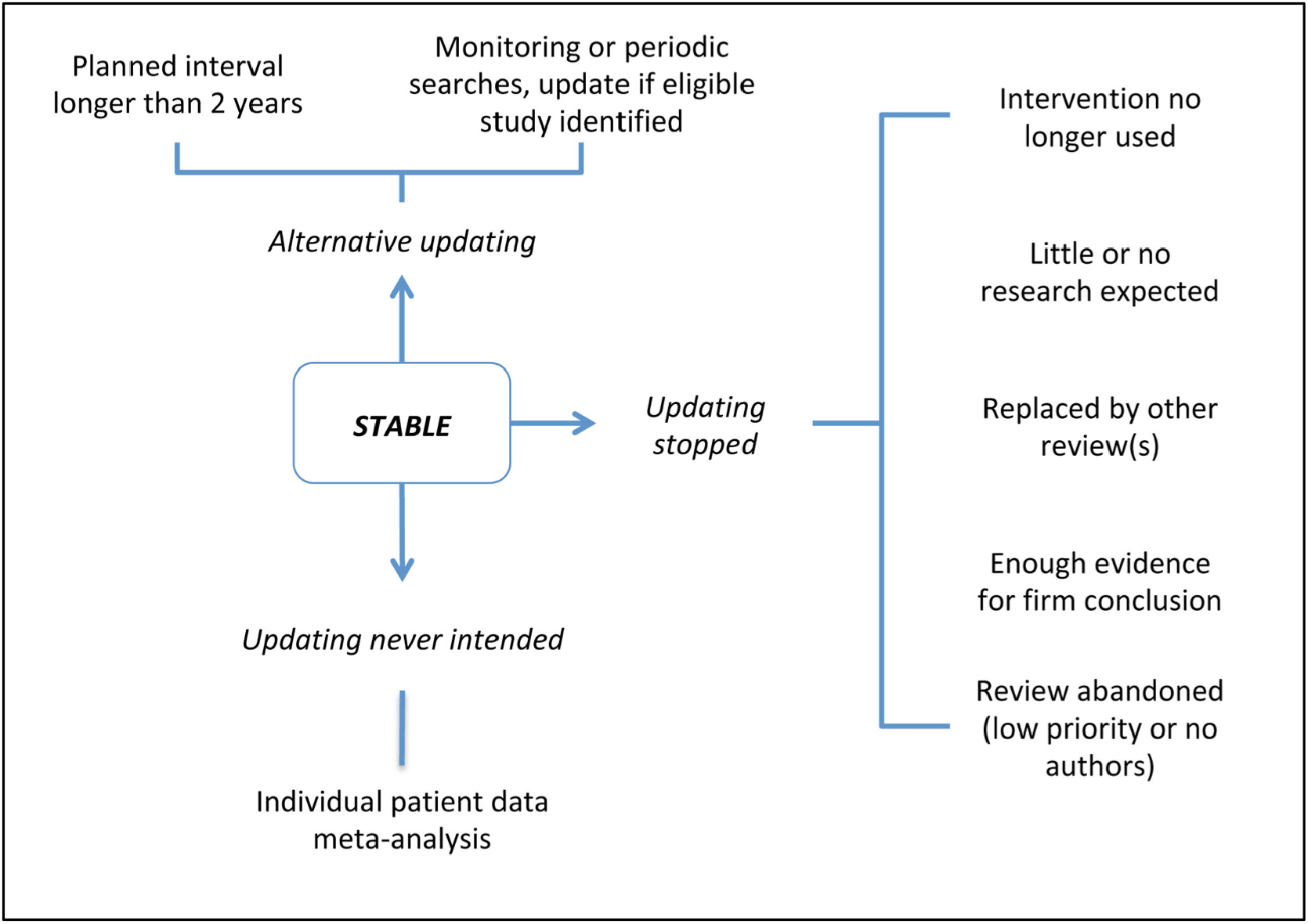
Categories of reasons reported for stable status of reviews.

Although each category is distinct, they are not mutually exclusive: a review could be assigned to ongoing monitoring rather than scheduled updating because little further research is expected, for example. Table 3 shows the proportion of reviews assigned to each of the eight categories, in order of frequency. For two reviews, no reasons were reported for declaring the review stable.

**Table 3.** Reported reasons for declaring Cochrane reviews stable (n = 505).

| Reason (ID) | Number | Percent |
| --- | --- | --- |
| Little or no further evidence expected (B) | 147 | 29.1 |
| Monitoring new trials or periodic searches, to be updated only if new eligible study identified or standards change (E) | 99 | 19.6 |
| Update required, but in more than 2 years (D) | 85 | 16.8 |
| Intervention not in general use, superseded, or withdrawn from market (A) | 60 | 11.9 |
| Review has been or will be superseded by a new review (F) | 49 | 9.7 |
| Firm conclusions; new studies would be unlikely to change conclusions (C) | 39 | 7.7 |
| Abandoned; low priority and/or authors unavailable for updating (G) | 21 | 4.2 |
| Individual patient data review (H) | 5 | 1.0 |
| **Total** | **505** | **100** |

The three most common reasons for declaring a review stable all relate to no, little, or infrequent research activity expected (331 of 505; 65.5%). As such a large proportion of these reviews were from one CRG, Figure 4 illustrates the proportion of reviews in each of the eight categories as in Table 2, with and without its reviews. This suggests that editorial practices were variable between CRGs that applied stable status.

**Figure 4.**
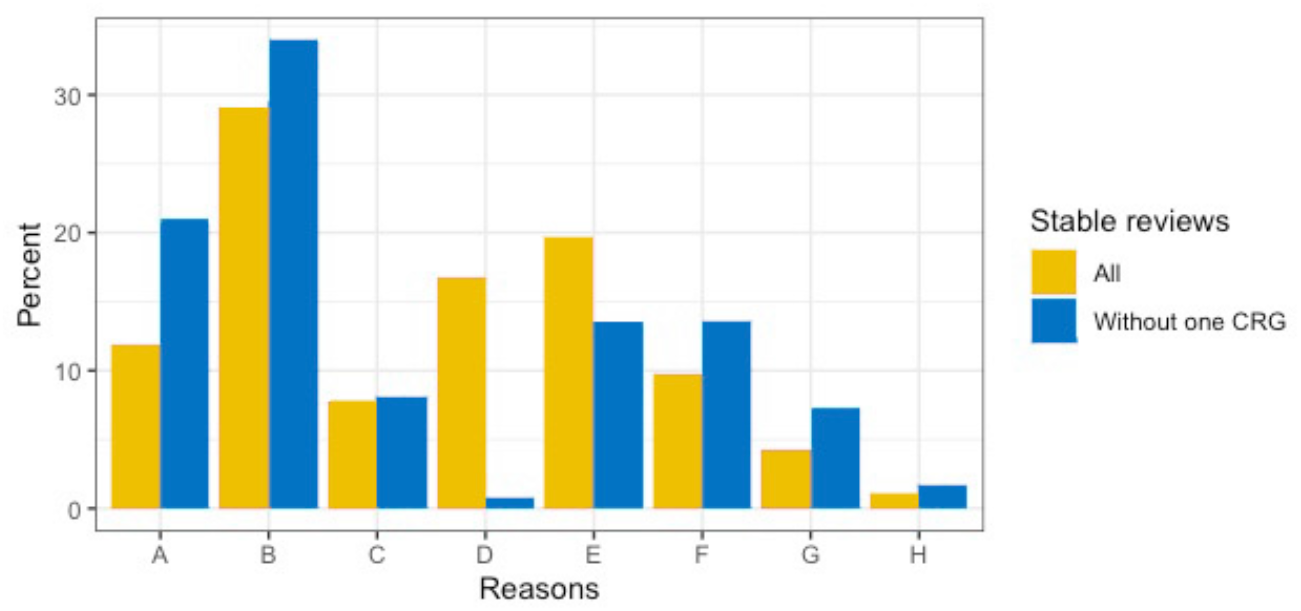
Reported reasons for declaring Cochrane reviews stable, with and without 219 reviews from a single CRG. Note: There were 505 stable reviews with reported reasons, and 286 without the 219 reviews from a single CRG.

### Reviews with firm conclusions reported to be unlikely to change

There were 39 reviews designated stable with firm conclusions reported to be unlikely to change with further evidence, which was 0.5% of all 7,645 non-withdrawn Cochrane reviews in 2019. They came from 11 CRGs, including 16 from the single CRG with the most stable reviews (41.0%, approximately the same proportion as the group has for stable reviews overall). Two of the reviews coming to firm conclusions reported an analytic method for this decision in their methods (5.1%). In one, it was cumulative meta-analysis, and in the other, it was a sample size calculation based on data from the larger included trials.

In 20 of the 39 reviews, the firm conclusion was that there was a benefit (51.3%). The firm conclusion in the 19 others was an absence of evidence of benefit or superiority, with five of those concluding there was evidence of adverse effect(s) (12.8%).

The majority of the reviews concluded there were still open questions. The authors in 20 of the 39 reviews with firm conclusions wrote that further research was needed on the subject for which their conclusion was firm (59.0%). Of the 16 reviews where authors concluded no further research on that question was needed, six recommended research on other questions related to the review’s subject (a further 15.4% of the 39 reviews).

Table 4 shows the evidence rating and future research recommendation for the 14 reviews that had GRADE SoF tables, broken down by the type of firm conclusion.

**Table 4.**
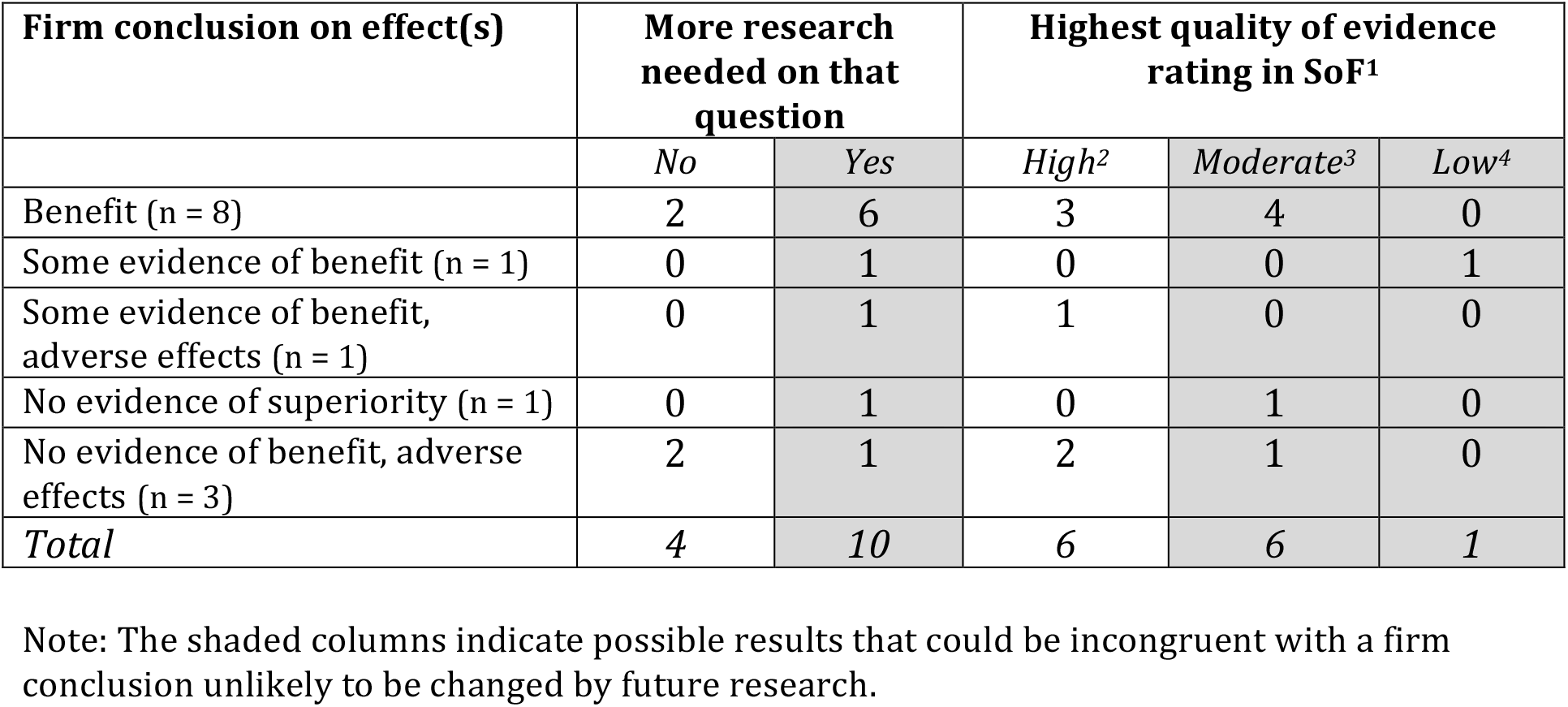
Reviews coming to firm conclusions, with a GRADE Summary of Findings (SoF) table (n = 14).

In those 14 reviews with evidence rated using GRADE, we saw little consistency between the firm conclusion/stable status, the quality of evidence, and the recommendation about future research. Only three of the 14 reviews (21.4%) rated the quality of the evidence as high, with no further research recommended. The others are not universally contradictory – for example, in one review, the lack of good evidence of effectiveness was coupled with reference to literature on the biological implausibility of possible benefit from the intervention. (37) That is exceptional, however, and the apparent internal contradictions in conclusions were typically not explained. Some of the discrepancy could be related to conflating the question of whether a strong enough study will be done, with what the impact of one would be.

1. One review with an SoF table did not rate the quality of the evidence.
2. Defined in the review as “High: further research is very unlikely to change our confidence in the estimate of effect”.
3. Defined in the review as “Moderate: further research is likely to have an important effect on our confidence in the estimate of effect and may change the estimate”.
4. Defined in the review as “Low: our confidence in the effect estimate is limited: the true effect may be substantially different from the estimate of the effect”.

For the group of 25 reviews without a GRADE SoF table, 12 concluded there was no need for further research on the subject of their firm conclusion (48.0%). However, we could not assess the consistency with the firm conclusion and the authors’ judgment of the strength of the evidence. Many reported no overall “rating” of the quality of the included evidence (14 of 25; 56.0%), and of those that did, there was no other consistent method for reaching that judgment.

### Use of analytic methods for determining the evidence is enough in protocols for Cochrane reviews

We identified 116 out of 2,415 review protocols that reported some planned use of one of the formal analytic methods we searched for (Table 5).

**Table 5.**
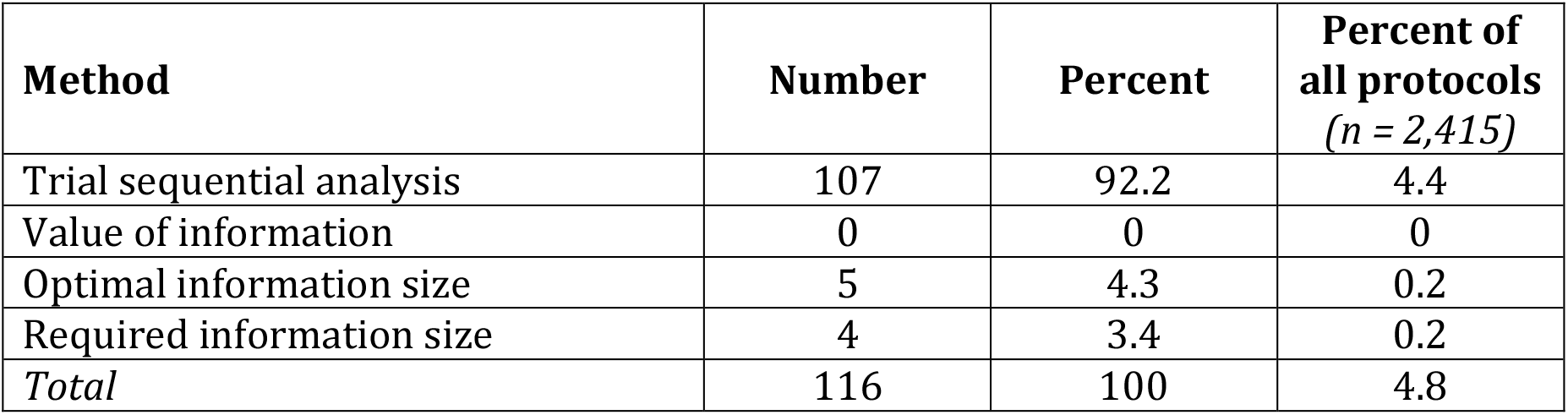
Use of analytic methods in protocols identified by text search (n = 116).

| Method | Number | Percent | Percent of all protocols<br>(*n* = 2,415) |
| --- | --- | --- | --- |
| Trial sequential analysis | 107 | 92.2 | 4.4 |
| Value of information | 0 | 0 | 0 |
| Optimal information size | 5 | 4.3 | 0.2 |
| Required information size | 4 | 3.4 | 0.2 |
| *Total* | 116 | 100 | 4.8 |

The method proposed was mostly trial sequential analysis, and most uses of it came from the CRG associated with the development of the method that advises its authors to use it (79 of 107, 73.8%). (38) Each use of optimal information size was in relation to GRADE assessment.

In all, 19 CRGs in this search had at least one protocol using one of these methods (35.8% of all CRGs). The range of protocols per CRG was 1 to 79 (median 1, IQR 2).

## Discussion

The group of reviews declared stable by Cochrane authors or editorial groups demonstrate several ways in which a “live” systematic review can reach its end. It was usually not because there was enough reliable evidence. There were some relatively common reasons for declaring a Cochrane review stable. One was that the research focus shifted or the review needed to be split or merged with another or others. Another was that the clinical question had lapsed because the interventions involved are no longer available or have been superseded by other forms of care.

However, the overwhelming reason for declaring reviews stable related to the perceived likelihood of there being further eligible studies. Two-thirds of the reviews were declared stable because of some variation of infrequent or no research activity around the intervention(s). This is also a critical factor to others. For example, for the National Institute for Health and Care Evidence (NICE), not identifying any major ongoing studies is key to retiring a question. (39)

Low likely research yield is a logical criterion for the use of scarce updating resources. However, following active research areas only could tilt the systematic review agenda towards the agendas of those who invest in trials, rather than clinical and consumer relevance. “Continuing importance of the review question to decision makers” is explicitly a key consideration about updating in Cochrane guidance. (10) That goal might be at risk from considering whether there are new studies that could impact results before deciding to update. The latest issue of the Cochrane Handbook also recommends considering metrics such as review clicks and citations in deciding whether or not to update. (10) That may lead in the same direction, perhaps channeling resources issues of less health value because they can go viral.

This situation becomes critical when the question still concerns consumers and clinicians and an updated systematic review could have encouraged further studies, or the judgment about the likelihood of important evidence proves to be wrong. In our sample, authors of at least 5% of stable reviews reversed their decision between 2013 and 2019, with two of those reviews having changed conclusions because of new evidence. Although that is reassuringly low, we do not know whether important new evidence or other developments affected further reviews that did not search for them.

The 2016 consensus statement on updating systematic reviews (14) argues that decisions not to update a systematic review need to be made in a context where new studies are under surveillance, because “… it is still important to assess new studies that might meet the inclusion criteria. New studies can show unexpected effects (eg, attenuation of efficacy) or provide new information about the effects seen in different circumstances (eg, groups of patients or locations)”. The remit of Cochrane’s review groups includes maintaining a register of trials within their subject scope, but we do not know the extent to which they all systematically assess incoming studies.

New studies are not the only development that could affect or compromise a systematic review’s conclusions and estimates. Identification of major error in an included study, for example, could reduce the effect size in a meta-analysis, (40) or could invalidate a review’s conclusions. The 2019 *Cochrane Handbook for Systematic Reviews of Interventions* notes studies retracted for data fabrication should be removed from Cochrane reviews. (10)

The designation “stable” has been replaced in 2019 by the status, “no update planned”. Given the problems our study shows with the implementation of the stable status, its end is justified. The replacement status has five options, none of which cover having enough evidence: (a) the intervention or (b) review is superseded, (c) the research area is no longer active, (d) the review is of low priority, and (e) “other”. (28)

“No update planned” is one of three potential statuses from 2019, the others being “up to date” and “update pending”. These categories aim to “provide readers with a guide to the status of the Cochrane review, and the likely future plans for the Cochrane Review with respect to updating”. (28) The usefulness of the “up to date” categorisation to readers depends on how current and accurate that judgment is. “Update pending” may be inherently misleading to readers, however. It could mean an update is around the corner, but it could be a euphemism for “out of date”.

Our study’s results have only limited application to Cochrane’s new system. Analysing these reviews was useful for charting part of the life cycle of systematic reviews and exploring some practices, but it had had major limitations in assessing prevalence of those practices. A substantial number of Cochrane’s editorial groups never applied the “stable” status, and practices among those that did appears to be highly variable. The data therefore do not reflect the proportion of Cochrane reviews to which the status could apply. Reasons for declaring a review stable were often poorly reported or ambiguously stated. A substantial proportion of our category assignments were based on internal Cochrane information without assessing the text of the review, or were made by a single author.

However, several of our findings are relevant for Cochrane reviews and for the development of methods in systematic reviews generally. We found that judgments about the conclusiveness of evidence and potential importance of future research to current findings were often inconsistent within the small group of reviews coming to firm conclusions. The potential for an error in judgment is high for Cochrane reviews, (21,41–43) and there can be considerable differences in authors’ GRADE-based assessments in systematic reviews in general. (44) Differing interpretations at the additional level suggested by this study underscore the value that improved methodology could offer.

Although the *Cochrane Handbook* discourages the use of methods such as trial sequential analysis, (10) value of information and related methods have been advocated or are in use for determining future research needs based on meta-analysis, (45,46) as well as methods for determining priorities in updating them. (14,47,48) We found that a small proportion of Cochrane protocols are incorporating similar methods to determine when updating is no longer required. Our search terms would not have identified all the protocols using a methodology to prespecify when a review could be closed.

Optimal information size is now explicitly recommended in the *Cochrane Handbook* for considering the imprecision of trial results, (10) and that may be more widely used in future. The impact of that should be assessed, both for systematic reviewers and users of reviews. The inconsistencies we identified in this study underscore the need for reliable analytic methods to support decision-making about the conclusiveness of evidence. Those decisions include health care choices and recommendations, as well as conducting, funding, approving, and participating in clinical trials. Being able to decide when there is enough evidence, with reasonable reliability, is both a practical and ethical necessity.

## Data Availability

Data and supporting files, including analytic code, are available on GitHub.

https://github.com/hildabast/enough-evidence

## Author contributions

HB initiated the study, curated, analysed and visualised data, and drafted the manuscript. Both authors contributed to the design of the study, study assessments, and interpretation of the data. HB acquired the data from the *CDSR* and Cochrane website, and LGH acquired the data from Archie. LGH revised the manuscript critically.

## Disclosures

Hilda Bastian was a member of the Cochrane Collaboration’s governing body from the organisation’s founding in 1993 until 2001, the coordinating editor of a CRG (Consumers and Communication) from 1997 to 2001, and a member of the GRADE working group from 2006 to 2012. She received support from an Australian Government Research Training Program Scholarship. Lars G. Hemkens is a member of the Cochrane Collaboration’s Adverse Events Methods Group and a CRG (Heart), and is co-author of several Cochrane reviews. Both authors previously worked for another organisation that undertakes systematic reviews, the German Institute for Quality and Efficiency in Healthcare (IQWiG).

## Acknowledgments

We are grateful to Toby Lasserson from Cochrane’s Editorial Unit for assistance with data on non-withdrawn reviews, and Anne Eisinga from the UK Cochrane Centre for assistance with a source on Cochrane’s original updating policy. We also thank the Cochrane Editorial Unit for guidance on the use of data from Archie.

## Abbreviations

CDSR: *Cochrane Database of Systematic Reviews* (journal component of The Cochrane Library)
CRG: Cochrane review group
GRADE: Grading of Recommendations, Assessment, Development and Evaluation
NICE: National Institute for Health and Care Evidence
SoF: GRADE Summary of Findings table in Cochrane reviews
XML: eXtensible Markup Language (for encoding human- and machine-readable files)

## Notes

### Competing Interest Statement

The authors have declared no competing interest.

### Funding Statement

No external funding was received. Hilda Bastian received support from an Australian Government Research Training Program Scholarship.

## References

1. Antman EM, Lau J, Kupelnick B, Mosteller F, Chalmers TC. A comparison of results of meta-analyses of randomized control trials and recommendations of clinical experts. Treatments for myocardial infarction. JAMA. 1992 Jul 8;268(2):240–8.

2. Chalmers I, Glasziou P. Avoidable waste in the production and reporting of research evidence. Lancet Lond Engl. 2009 Jul 4;374(9683):86–9.

3. Moher D, Stewart L, Shekelle P. All in the Family: systematic reviews, rapid reviews, scoping reviews, realist reviews, and more. Syst Rev. 2015 Dec 22;4(1):183.

4. Shojania KG, Sampson M, Ansari MT, Ji J, Doucette S, Moher D. How quickly do systematic reviews go out of date? A survival analysis. Ann Intern Med. 2007 Aug 21;147(4):224–33.

5. Chalmers I, Hedges LV, Cooper H. A Brief History of Research Synthesis. Eval Health Prof. 2002 Mar 1;25(1):12–37.

6. Chalmers I. The Cochrane Collaboration: Preparing, Maintaining, and Disseminating Systematic Reviews of the Effects of Health Care. Ann N Y Acad Sci. 1993;703(1):156–65.

7. Starr M, Chalmers I, Clarke M, Oxman AD. The origins, evolution, and future of The Cochrane Database of Systematic Reviews. Int J Technol Assess Health Care. 2009 Jul;25 Suppl 1:182–95.

8. Chandler J, Hopewell S. Cochrane methods - twenty years experience in developing systematic review methods. Syst Rev. 2013 Sep 20;2(1):76.

9. Cochrane Collaboration. Minutes of meeting of the Cochrane Collaboration Steering Group, Cape Town, South Africa, 24 and 29 October 2000 [Internet]. Cochrane Collaboration; 2000. Available from: https://community.cochrane.org/sites/default/files/uploads/inline-files/minutes_reports/ccsg/2000.10.24.htm

10. Higgins JPT, Thomas J. Cochrane Handbook for Systematic Reviews of Interventions [Internet]. Version 6.0. London: Cochrane Collaboration; 2019 [cited 2019 Oct 2]. Available from: https://training.cochrane.org/handbook/current

11. Bastian H, Glasziou P, Chalmers I. Seventy-Five Trials and Eleven Systematic Reviews a Day: How Will We Ever Keep Up? PLOS Med. 2010 Sep 21;7(9):e1000326.

12. Garritty C, Tsertsvadze A, Tricco AC, Sampson M, Moher D. Updating systematic reviews: an international survey. PloS One. 2010 Apr 1;5(4):e9914.

13. Tsertsvadze A, Maglione M, Chou R, Garritty C, Coleman C, Lux L, et al. Updating Comparative Effectiveness Reviews: Current Efforts in AHRQ’s Effective Health Care Program. In: Methods Guide for Effectiveness and Comparative Effectiveness Reviews [Internet]. Rockville (MD): Agency for Healthcare Research and Quality (US); 2011 [cited 2019 Sep 16]. (AHRQ Methods for Effective Health Care). Available from: http://www.ncbi.nlm.nih.gov/books/NBK66066/

14. Garner P, Hopewell S, Chandler J, MacLehose H, Schünemann HJ, Akl EA, et al. When and how to update systematic reviews: consensus and checklist. BMJ. 2016 20;354:i3507.

15. Egger M, Smith GD. Misleading meta-analysis. BMJ. 1995 Mar 25;310(6982):752–4.

16. Pogue JM, Yusuf S. Cumulating evidence from randomized trials: utilizing sequential monitoring boundaries for cumulative meta-analysis. Control Clin Trials. 1997 Dec;18(6):580–93; discussion 661-666.

17. Pogue J, Yusuf S. Overcoming the limitations of current meta-analysis of randomised controlled trials. Lancet Lond Engl. 1998 Jan 3;351(9095):47–52.

18. Egger M, Smith GD, Sterne JA. Meta-analysis: is moving the goal post the answer? Lancet Lond Engl. 1998 May 16;351(9114):1517.

19. Nüesch E, Jüni P. Commentary: Which meta-analyses are conclusive? Int J Epidemiol. 2009 Feb;38(1):298–303.

20. Wetterslev J, Thorlund K, Brok J, Gluud C. Trial sequential analysis may establish when firm evidence is reached in cumulative meta-analysis. J Clin Epidemiol. 2008 Jan;61(1):64–75.

21. Brok J, Thorlund K, Wetterslev J, Gluud C. Apparently conclusive meta-analyses may be inconclusive--Trial sequential analysis adjustment of random error risk due to repetitive testing of accumulating data in apparently conclusive neonatal meta-analyses. Int J Epidemiol. 2009 Feb;38(1):287–98.

22. Imberger G, Thorlund K, Gluud C, Wetterslev J. False-positive findings in Cochrane meta-analyses with and without application of trial sequential analysis: an empirical review. BMJ Open. 2016 12;6(8):e011890.

23. Cochrane Collaboration. Minutes of meetings of the Cochrane Collaboration Steering Group, Khon Kaen, Thailand 24-26 April 2006 [Internet]. Cochrane Collaboration; 2006 [cited 2019 Sep 20]. Available from: https://community.cochrane.org/sites/default/files/uploads/inline-files/minutes_reports/ccsg/1MinutesofKhonKaenCCSGmeetingwithposthoccorrections.htm

24. Higgins JPT, Green SE. Cochrane Handbook for Systematic Reviews of Interventions [Internet]. Version 5.0.0. Oxford: Cochrane Collaboration; 2008. Available from: https://handbook-5-1.cochrane.org/v5.0.0/

25. Guyatt GH, Oxman AD, Vist GE, Kunz R, Falck-Ytter Y, Alonso-Coello P, et al. GRADE: an emerging consensus on rating quality of evidence and strength of recommendations. BMJ. 2008 Apr 26;336(7650):924–6.

26. Hultcrantz M, Rind D, Akl EA, Treweek S, Mustafa RA, Iorio A, et al. The GRADE Working Group clarifies the construct of certainty of evidence. J Clin Epidemiol. 2017 Jul;87:4–13.

27. Schünemann H, Brożek J, Guyatt G, Oxman AD. GRADE Handbook [Internet]. Hamilton: GRADE; 2013. Available from: https://gdt.gradepro.org/app/handbook/handbook.html

28. Cochrane Editorial Unit. Updating Classification System: guide to applying to Cochrane Reviews [Internet]. Cochrane Collaboration; 2019. Available from: https://community.cochrane.org/sites/default/files/uploads/inline-files/Cochrane_UCS-Guide_8Jul16_0.pdf#targetText=The%20Updating%20Classification%20System%20(UCS,decisions%20for%20individual%20Cochrane%20Reviews.

29. Ahtirschi O, van Valkenhoef G. Advanced document search options - Archie Help [Internet]. Documentation.cochrane.org. 2019 [cited 2019 Sep 23]. Available from: https://documentation.cochrane.org/display/Archie/Advanced+document+search+options

30. Archie [Internet]. London: Cochrane Collaboration; 2019. Available from: https://community.cochrane.org/help/tools-and-software/archie

31. Cochrane Collaboration. Cochrane Review Groups and Networks [Internet]. Cochrane Library. [cited 2019 Oct 10]. Available from: https://www.cochranelibrary.com/about/cochrane-review-groups

32. RStudio Team. RStudio: integrated development environment for R. [Internet]. Boston: RStudio Inc; 2015 [cited 2019 Aug 12]. Available from: https://www.rstudio.com/

33. R Core Team. R: a language and environment for statistical computing [Internet]. Vienna: R Foundation for Statistical Computing; 2016. Available from: https://www.r-project.org/

34. Wickham H. tidyverse [Internet]. RStudio Inc; 2017. Available from: https://CRAN.R-project.org/package=tidyverse

35. Wickham H. reshape2 [Internet]. 2017. Available from: https://CRAN.R-project.org/package=reshape2

36. Bastian H. Enough evidence. GitHub [Internet]. 2019 Nov; Available from: https://github.com/hildabast/enough-evidence

37. Enayati AA, Hemingway J, Garner P. Electronic mosquito repellents for preventing mosquito bites and malaria infection. Cochrane Database Syst Rev. 2007 Apr 18;(2):CD005434.

38. Cochrane Hepato-Biliary. Information for authors [Internet]. Cochrane Hepato-Biliary. [cited 2019 Sep 29]. Available from: /information-authors

39. National Institute for Health and Care Excellence. Developing NICE Guidelines: The Manual [Internet]. London: National Institute for Health and Care Excellence (NICE); 2015 [cited 2019 Aug 24]. (NICE Process and Methods Guides). Available from: http://www.ncbi.nlm.nih.gov/books/NBK310375/

40. Fanelli D, Moher D. What difference do retractions make? An estimate of the epistemic impact of retractions on recent meta-analyses [Internet]. bioRxiv; 2019. Available from: https://www.biorxiv.org/content/10.1101/734137v1

41. Fleming PS, Koletsi D, Ioannidis JPA, Pandis N. High quality of the evidence for medical and other health-related interventions was uncommon in Cochrane systematic reviews. J Clin Epidemiol. 2016;78:34–42.

42. Garcia-Alamino JM, Bankhead C, Heneghan C, Pidduck N, Perera R. Impact of heterogeneity and effect size on the estimation of the optimal information size: analysis of recently published meta-analyses. BMJ Open. 2017 Nov 8;7(11):e015888.

43. Gartlehner G, Nussbaumer-Streit B, Wagner G, Patel S, Swinson-Evans T, Dobrescu A, et al. Increased risks for random errors are common in outcomes graded as high certainty of evidence. J Clin Epidemiol. 2019 Feb;106:50–9.

44. Berkman ND, Lohr KN, Morgan LC, Kuo T-M, Morton SC. Interrater reliability of grading strength of evidence varies with the complexity of the evidence in systematic reviews. J Clin Epidemiol. 2013 Oct;66(10):1105-1117.e1.

45. Myers E, Sanders GD, Ravi D, Matchar D, Havrilesky L, Samsa G, et al. Evaluating the Potential Use of Modeling and Value-of-Information Analysis for Future Research Prioritization Within the Evidence-Based Practice Center Program [Internet]. Rockville (MD): Agency for Healthcare Research and Quality (US); 2011 [cited 2019 Sep 22]. (AHRQ Methods for Effective Health Care). Available from: http://www.ncbi.nlm.nih.gov/books/NBK62134/

46. Roloff V, Higgins JPT, Sutton AJ. Planning future studies based on the conditional power of a meta-analysis. Stat Med. 2013 Jan 15;32(1):11–24.

47. Sampson M, Shojania KG, McGowan J, Daniel R, Rader T, Iansavichene AE, et al. Surveillance search techniques identified the need to update systematic reviews. J Clin Epidemiol. 2008 Aug;61(8):755–62.

48. Sutton AJ, Donegan S, Takwoingi Y, Garner P, Gamble C, Donald A. An encouraging assessment of methods to inform priorities for updating systematic reviews. J Clin Epidemiol. 2009 Mar;62(3):241–51.

